# Associations between different tau-PET patterns and longitudinal atrophy in the Alzheimer’s disease continuum

**DOI:** 10.1101/2021.08.10.21261824

**Authors:** Rosaleena Mohanty, Daniel Ferreira, Agneta Nordberg, Eric Westman, the Alzheimer’s Disease Neuroimaging Initiative

## Abstract

**INTRODUCTION:** Different subtypes/patterns have been defined using tau-PET and structural-MRI in Alzheimer’s disease (AD), but the relationship between tau pathology and atrophy remains unclear. Our goals were twofold: (a) investigate the association between baseline tau-PET patterns and longitudinal atrophy in the AD continuum; (b) characterize *heterogeneity* as a continuous phenomenon over the conventional notion using discrete subgroups.

**METHODS:** In 366 individuals (amyloid-beta-positive: cognitively normal, prodromal AD, AD dementia; amyloid-beta-negative healthy), we examined the association between tau-PET patterns (operationalized as a continuous phenomenon and a discrete phenomenon) and longitudinal sMRI.

**RESULTS:** We observed a differential association between tau-PET patterns and longitudinal atrophy. Heterogeneity, measured continuously, may offer an alternative characterization, sharing correspondence with the conventional subgrouping.

**DISCUSSION:** Site and the rate of atrophy are modulated differentially by tau-PET patterns in the AD continuum. We postulate that *heterogeneity* be treated as a continuous phenomenon for greater sensitivity over the current/conventional discrete subgrouping.

## 1. Introduction

The biological framework of Alzheimer’s disease (AD) recognizes beta-amyloid (Aβ), tau and neurodegeneration as the characteristic biomarkers in disease pathogenesis[1]. Among the AD hallmarks, spread of Aβ in the brain is rather diffuse whereas tau accumulates relatively locally and gradually[2]. Occurrence of neurodegeneration downstream to Aβ and tau has incited several investigations on the relationship among these biomarkers, suggesting a closer association between neurodegeneration and tau than neurodegeneration and Aβ[3–5].

Biological heterogeneity in AD manifests as distinct patterns in the pre-dementia stages, which likely evolve into subtypes in the dementia stage. Neuroimaging studies have shown topographical conformity and association between tau pathology from tau positron emission tomography (tau-PET) and longitudinal brain atrophy-based neurodegeneration from magnetic resonance imaging (MRI), in cognitively unimpaired individuals[6], prodromal AD and/or AD dementia[4,7,8] and clinical subtypes of AD[9,10]. A critical caveat, however, is the failure to account for heterogeneity in tau-PET topography at a given disease stage (i.e., tau patterns or subtypes)[11–15]. The relationship between tau-PET patterns and atrophy remains unexplored and is important for precision medicine.

We primarily investigated the association between different tau-PET patterns and longitudinal atrophy in the AD continuum (cognitively normal, prodromal AD, AD dementia with Aβ pathology). We secondarily characterized tau-PET patterns on a continuous-scale inspired by the recent conceptual framework[16], compared to and extending beyond the conventional characterization of discrete categorization[13– 15,17]. We hypothesized that (a) tau-PET patterns would modulate the association between baseline tau-PET and longitudinal atrophy differentially; (b) treating *heterogeneity* (the different tau-PET patterns) on a continuous-scale over a discrete-scale could be more appropriate.

## 2. Methods

### 2.1. Participants

Participants were chosen from the Alzheimer’s disease neuroimaging initiative (ADNI; launched in 2003; PI: Michael W. Weiner; http://adni.loni.usc.edu/), aimed at measuring the progression of prodromal, early AD using biomarkers and clinical and neuropsychological assessments. We included 366 individuals including 173 Aβ+ individuals in the AD continuum (98 cognitively normal, 50 prodromal AD, 25 AD dementia) and 193 Aβ-cognitively normal individuals. All individuals had tau-PET and MRI cross-sectionally (*baseline*). Longitudinal MRI was available *retrospectively* (N=167) and *prospectively* (N=178). The ADNI protocol was approved by the local institutional review boards and all participants provided written informed consent (additional details in **Supplementary Section 1**).

### 2.2. Neuroimaging

#### MRI

MRI, collected on 3.0 T scanners, were 3-D accelerated T1-weighted sequences acquired sagittally with voxel size 1.1×1.1×1.2mm^3^. MRI processing is described in **Supplementary Section 2**. Regional thickness in 68 brain areas[18] represented brain atrophy.

#### Tau-PET

Tau-PET were collected on PET/CT scanners. [^18^F]AV-1451 was injected with a dosage of 370 MBq (10.0 mCi)±10% and scans were acquired between 75-105 min post-injection. Dynamic acquisition was 30 min long with 6×5 min frames. MRI was available within 90 days (except in 5 prodromal AD and 3 AD dementia patients, >90 days). Tau-PET processing is described in **Supplementary Section 2**. Regional tau-PET signal in the same 68 brain areas as MRI was quantified as standardized uptake value ratio (SUVR).

### 2.3. Regions of interest

We examined regional tau-PET SUVR and thickness, averaged bilaterally, in medial temporal and association cortical areas as consistently identified across tau-PET/MRI-based subtyping studies in AD[14]. More specifically, medial temporal lobe was represented by the *entorhinal cortex* unless specified otherwise. Hippocampus was excluded as it may suffer from off-target binding with the current tau-PET tracer[19,20]. *Neocortex* included the middle frontal, inferior parietal and superior frontal regions unless specified otherwise. Global tau-PET SUVR was calculated by averaging across all cortical/subcortical regions (except hippocampus).

### 2.4. Characterization of tau-PET patterns

We investigated *heterogeneous tau-PET patterns* using two different characterizations of baseline tau-PET:

#### 2.4.1. Tau-PET patterns on a continuous-scale

We followed the recently proposed conceptual framework for AD subtypes[16], quantifying two dimensions of tau-PET patterns in our cohort, measured on a continuous-scale: *typicality*, proxied by the ratio of entorhinal tau-PET SUVR to neocortical tau-PET SUVR (hereon referred to as *E:N*), similar to the index in the original neuropathological study[11]; and *severity*, proxied by the global tau-PET SUVR.

#### 2.4.2. Tau-PET patterns on a discrete-scale

We translated a MRI-based subtyping method[21] to tau-PET, using the healthy (Aβ-) reference group to characterize patterns within our target population without assumptions on the within-population distribution (**Supplementary Section 3**). This method compares entorhinal and neocortical SUVR and assigns individuals into one of four discrete patterns: *typical AD, limbic predominant, cortical predominant* or *minimal tau*. We have previously reported how this operationalization relates to other discrete-scale operationalizations[14].

### 2.5. Statistical analysis

We compared the groups within the AD continuum and the healthy individuals by demographics and clinical variables[22,23] using hypothesis testing (Kruskal–Wallis test for the continuous variables; Fisher exact test for the nominal variables).

We investigated our primary aim of association between tau-PET SUVR and longitudinal atrophy for the two characterizations of tau-PET patterns:

#### 2.5.1. Association between baseline tau-PET patterns (continuous-scale) and longitudinal atrophy

We assessed the relationship of tau-PET patterns, characterized by typicality (E:N) and severity (global tau-PET SUVR) measured on a continuous-scale, to regional thickness changes with linear regression models (**Supplementary Section 4**). Testing our primary hypothesis that tau-PET patterns may be differentially associated with longitudinal atrophy, we modeled tau-PET patterns (typicality and severity) and estimated regional thickness changes using linear mixed effects model (**Supplementary Section 4**). The main effects included the interaction between time and each of typicality and severity. For visualization, we stratified by typicality, assessing atrophy changes across the two patterns of this dimension (limbic predominance versus cortical predominance). Similarly, we stratified by severity, comparing atrophy changes across the two patterns of this dimension (typical AD versus minimal tau). Per stratification, we computed the annualized thickness change (%/year) across individuals for retrospective-to-baseline and baseline-to-prospective changes.

#### 2.5.2. Association between baseline tau-PET patterns (discrete-scale) and longitudinal atrophy

Testing our primary hypothesis that tau-PET patterns may be differentially associated with longitudinal atrophy, we also examined the association between tau-PET SUVR and longitudinal regional thickness changes within typical AD, limbic predominant, cortical predominant and minimal tau patterns using linear regression models controlled for the interval of longitudinal MRI. We assessed the full cohort (baseline N=173) and a subcohort, tracking the same set of individuals with MRI across all timepoints (N=61).

Analyses were performed using MATLAB R2020b (The MathWorks, Inc., Natick, Massachusetts, USA). Brain visualizations were generated through R v4.0.3 with ggseg (https://lcbc-uio.github.io/ggseg/).

### 2.6. A/T/longitudinal-N classification of tau-PET patterns

For a deeper understanding of tau-PET patterns, we analyzed the A/T/longitudinal-N (Aβ/Tau/longitudinal-atrophy-based Neurodegeneration) biomarker scheme[1] across them. We dichotomized each biomarker evaluating: Aβ positivity with global amyloid-PET SUVR (A+), tau positivity with regional tau-PET-based SUVR (**Supplementary Section 5**; T+) and neurodegeneration positivity at retrospective, baseline and prospective timepoints (N_R_+, N_B_+, N_P_+) with regional MRI-based thickness/atrophy (**Supplementary Section 6**). In the main report, we include tau and neurodegeneration positivity assessed in the entorhinal cortex and the neocortex. We compared the proportion (%) of A/T/longitudinal-N positivity across tau-PET patterns.

## 3. RESULTS

### 3.1. Participants

Clinical groups in our cohort were significantly different in age, education, *APOE* ε4 carriers and global cognition **(Supplementary Table 1)**. For longitudinal MRI, the retrospective-to-baseline time interval was 2.2±1years while the baseline-to-prospective time interval was 1.3±0.4years.

### 3.2. Characterization of tau-PET patterns

#### 3.2.1. Tau-PET patterns on a continuous-scale

**Figure 1A-B** shows the distribution of tau-PET patterns, characterized by typicality (E:N) and severity (global tau-PET SUVR) on a continuous-scale. Within the AD continuum, both dimensions showed the lowest variance in the Aβ+ cognitively normal (σ^2^=0.07 for typicality, σ^2^=0.05 for severity), an intermediate variance in prodromal AD (σ^2^=0.09 for typicality, σ^2^=0.1 for severity) and the highest variance in AD dementia (σ^2^=0.16 for typicality, σ^2^=0.52 for severity) (**Figure 1B**). There was no significant association between typicality and severity (r=0.003, *p*=0.97) in the AD continuum.

**Figure 1.**
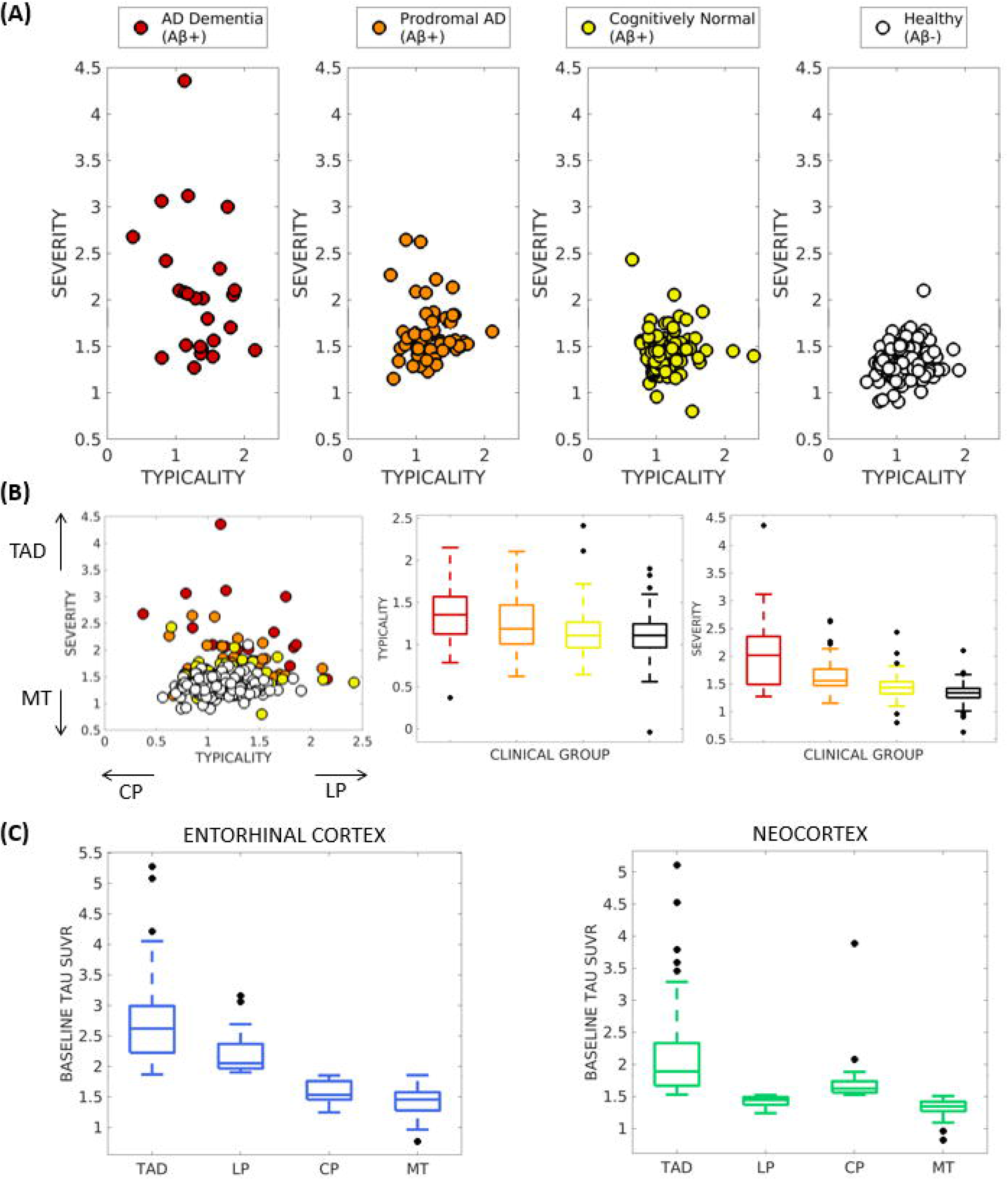
Baseline tau-PET patterns in the AD continuum characterized on a **(A, B) c**ontinuous scale and **(C)** discrete scale. Note: **(A)** Tau-PET patterns were assessed on a continuous scale across the AD continuum (including cognitively normal, prodromal AD and AD dementia), shown separately in each clinical group for visual comparison. **(B)** Tau-PET patterns assessed on a continuous scale, shown relative to the other clinical groups (left) with the relative tendency of typicality (in terms of CP or LP) and severity (in terms of TAD or MT) shown for correspondence with **(C)**; variability of typicality (center) and severity (right) across the clinical groups are presented. **(C)** Tau-PET patterns, assessed on a discrete scale in the AD continuum relative to the healthy group, identified four patterns whose tau-PET SUVR are shown in the entorhinal cortex (left) and the neocortex (right). AD = Alzheimer’s disease; TAD = typical AD pattern; LP = limbic predominant pattern; CP = cortical predominant pattern; MT = minimal tau.

#### 3.2.2. Tau-PET patterns on a discrete-scale

**Figure 1C** presents four discrete tau-PET patterns, based on the entorhinal and neocortical tau-PET SUVR: 33% typical AD pattern (N=57), 12% limbic predominant pattern (N=21), 18% cortical predominant pattern (N=31) and 37% minimal tau pattern (N=64). Demographic and clinical characteristics across patterns are summarized in **Supplementary Table 2**. The tau-PET patterns were significantly different in education, global and composite cognitive scores.

### 3.3. Association between tau-PET patterns and longitudinal atrophy

#### 3.3.1. Tau-PET patterns on a continuous-scale

**Supplementary Table 3** presents the association of baseline tau-PET patterns (typicality and severity) with longitudinal regional thickness change. We observed that: (i) individuals in the AD continuum (Aβ+), but not cognitively normal (Aβ-), showed significant associations of typicality and severity with longitudinal regional thickness changes; (ii) entorhinal cortex: typicality was associated with retrospective-to-baseline thinning while severity was associated with both retrospective-to-baseline and baseline-to-prospective thinning; (iii) neocortex: typicality was associated with retrospective-to-baseline thinning while severity was associated with baseline-to-prospective thinning; (iv) all significant associations were positive: for typicality, increased E:N (limbic predominant pattern) corresponded with greater regional thickness change while for severity, increased global tau-PET SUVR (typical AD pattern) corresponded with greater regional thickness change.

**Table 1** shows the estimated changes in longitudinal regional thickness using baseline tau-PET. Assessing our primary hypothesis of differential association of tau-PET patterns (typicality and severity) with longitudinal atrophy, the main effects included the interaction of each of typicality and severity with time on estimation of regional thickness change (% change/year).

**Table 1.** Estimated rate of atrophy change as a function of baseline tau-PET patterns (continuous-scale) in the AD continuum.

| Fixed Effects | Entorhinal cortex |  | Neocortex |  |
| --- | --- | --- | --- | --- |
|  | Estimate (SE) | *p* | Estimate (SE) | *p* |
| Intercept | **4.75 (0.36)** | **<0.0001** | **2.89 (0.08)** | **<0.0001** |
| Age | **-0.01 (0.003)** | **<0.0001** | **-0.004 (0.0009)** | **0.0002** |
| T<sub>R</sub> | **-0.38 (0.08)** | **<0.0001** | **-0.05 (0.015)** | **0.0009** |
| T<sub>P</sub> | **0.21 (0.07)** | **0.004** | **0.04 (0.014)** | **0.004** |
| Tau<sub>B</sub> | -0.24 (0.13) | 0.06 | **-0.08 (0.04)** | **0.042** |
| Typicality | 0.08 (0.19) | 0.67 | **-0.07 (0.03)** | **0.012** |
| Severity | -0.11 (0.18) | 0.56 | 0.019 (0.05) | 0.72 |
| Tau<sub>B</sub> × T<sub>R</sub> | -0.05 (0.04) | 0.19 | 0.02 (0.01) | 0.22 |
| Tau<sub>B</sub> × T<sub>P</sub> | -0.014 (0.04) | 0.69 | -0.03 (0.01) | 0.052 |
| Typicality<sub>B</sub> × T<sub>R</sub> | **0.21 (0.06)** | **0.0005** | **0.02 (0.01)** | **0.027** |
| Typicality<sub>B</sub> × T<sub>P</sub> | -0.08 (0.05) | 0.13 | -0.01 (0.01) | 0.31 |
| Severity<sub>B</sub> × T<sub>R</sub> | **0.17 (0.06)** | **0.003** | -0.005 (0.02) | 0.81 |
| Severity<sub>B</sub> × T<sub>P</sub> | -0.08 (0.05) | 0.09 | 0.01 (0.02) | 0.59 |

Changes in atrophy (thickness) are estimated by the linear mixed effects model with individual-specific intercepts. The linear mixed effects model was centered at T<sub>B</sub>. Longitudinal atrophy was modeled as the dependent variable. Age, time, Tau<sub>B</sub>, Typicality<sub>B</sub>, Severity<sub>B</sub> and interactions with time were modeled as fixed effects. The significant effects corresponding to $p \leq 0.05$ are reported in **bold**; SE = Standard error in coefficient; T<sub>R</sub> = retrospective timepoint; T<sub>B</sub> = baseline timepoint; T<sub>P</sub> = prospective timepoint, Tau<sub>B</sub> = baseline tau-PET SUVR, Typicality<sub>B</sub> = baseline typicality (proxied by E:N), Severity<sub>B</sub> = baseline severity (proxied by global tau-PET SUVR).

##### Typicality × Time interaction

**Table 1** shows that the estimated longitudinal thickness changes were significant for retrospective-to-baseline timepoints for the entorhinal cortex and neocortex. Stratifying by typicality (E:N) showed faster retrospective-to-baseline entorhinal thinning for higher E:N (limbic predominant pattern, -1.6%/year) than for lower E:N (cortical predominant pattern, +0.2%/year) (**Figure 2A**). Similarly retrospective-to-baseline neocortical thinning was faster for higher E:N (−0.3%/year) than for lower E:N (+0.2%/year) (**Figure 2A**). Baseline-to-prospective thickness changes relative to typicality were not significant.

**Figure 2.**
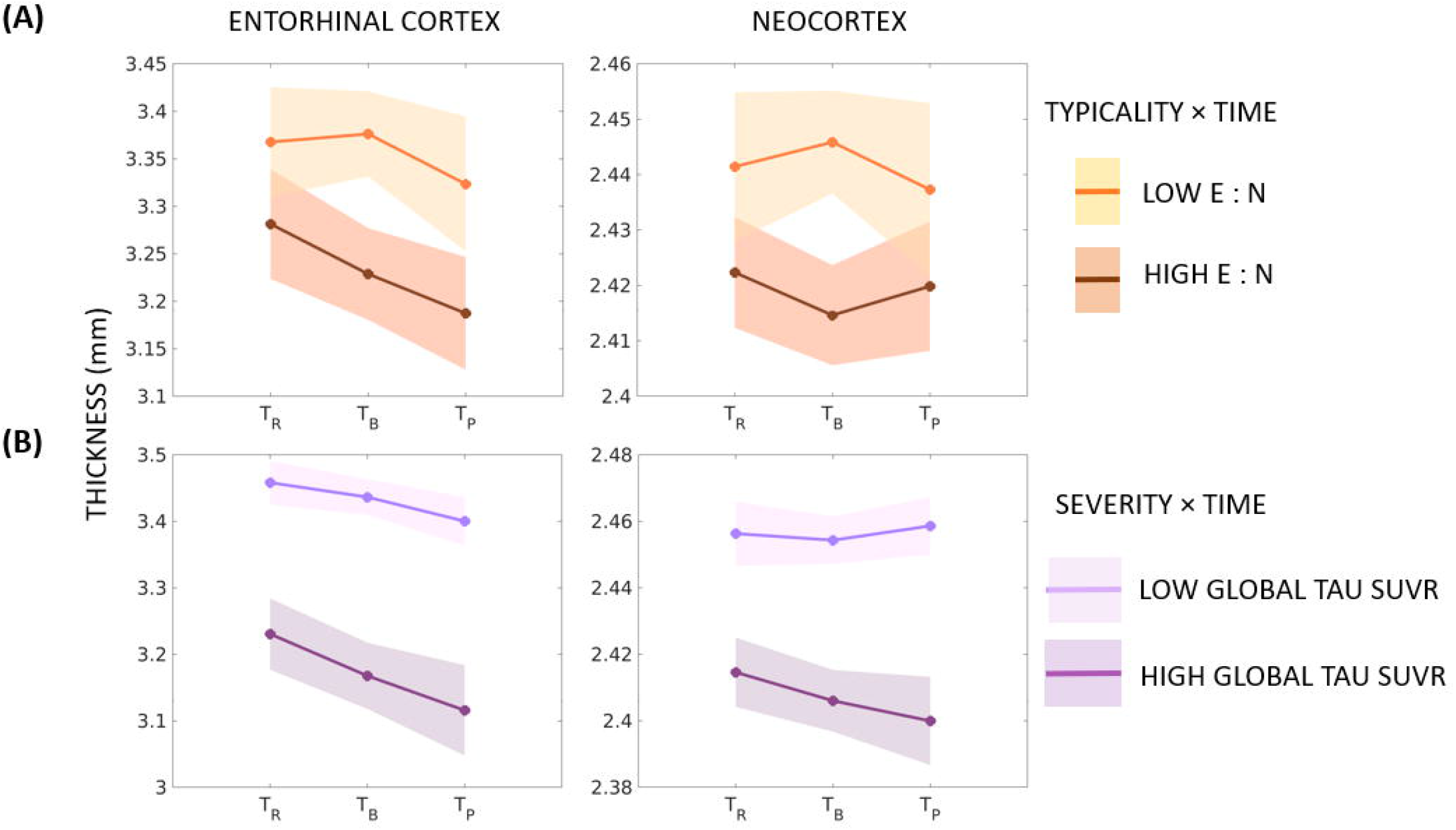
Longitudinal changes in atrophy relative to baseline tau-PET patterns (continuous scale) in the AD continuum. Note: Estimated longitudinal atrophy (thickness) estimated by linear mixed effects model for the entorhinal cortex and neocortex: **(A)** stratified by levels of typicality (the low/high groups were computed by median split in typicality); **(B)** stratified by levels of severity (the low/high groups were computed by median split in severity). Shaded regions represent the 95% confidence interval; T_R_ = retrospective timepoint; T_B_ = baseline timepoint; T_P_ = prospective timepoint; E:N = ratio of average entorhinal tau-PET SUVR to average neocortical tau-PET SUVR; SUVR = standardized uptake value ratio.

##### Severity × Time interaction

**Table 1** shows that the estimated longitudinal change in thickness was significant for retrospective-to-baseline timepoints for the entorhinal cortex. Stratifying by severity (global tau SUVR) showed faster retrospective-to-baseline entorhinal thinning for higher global tau SUVR (typical AD pattern, -1.9%/year) than for lower global tau SUVR (minimal tau pattern, -0.6%/year) (**Figure 2B**). Baseline-to-prospective thickness changes relative to severity were not significant.

#### 3.3.2. Tau-PET patterns on a discrete-scale

**Figure 3A** (top panel) shows the topography of the discrete tau-PET patterns at baseline (N=173): typical AD pattern had elevated tau-PET SUVR in the medial temporal lobe and in the remaining cortex; limbic predominant pattern had elevated tau-PET SUVR in the entorhinal cortex compared to the remaining cortex; cortical predominant pattern had elevated tau-PET SUVR in the cortex compared to the medial temporal lobe; minimal tau pattern did not show marked elevation of tau-PET SUVR in any region. **Figure 3A** (bottom panels) presents the longitudinal topography of regional thinning *within* each tau-PET pattern in a subcohort (N=61) of the AD continuum, tracked across all timepoints. Generally, regional thinning appeared pronounced at later timepoints. Topography of tau-PET SUVR elevation shared similarity with the topography of prospective regional thinning, mainly for typical AD and limbic predominant patterns. Clinical groups within each pattern are reported in **Supplementary Table 4**.

**Figure 3.**
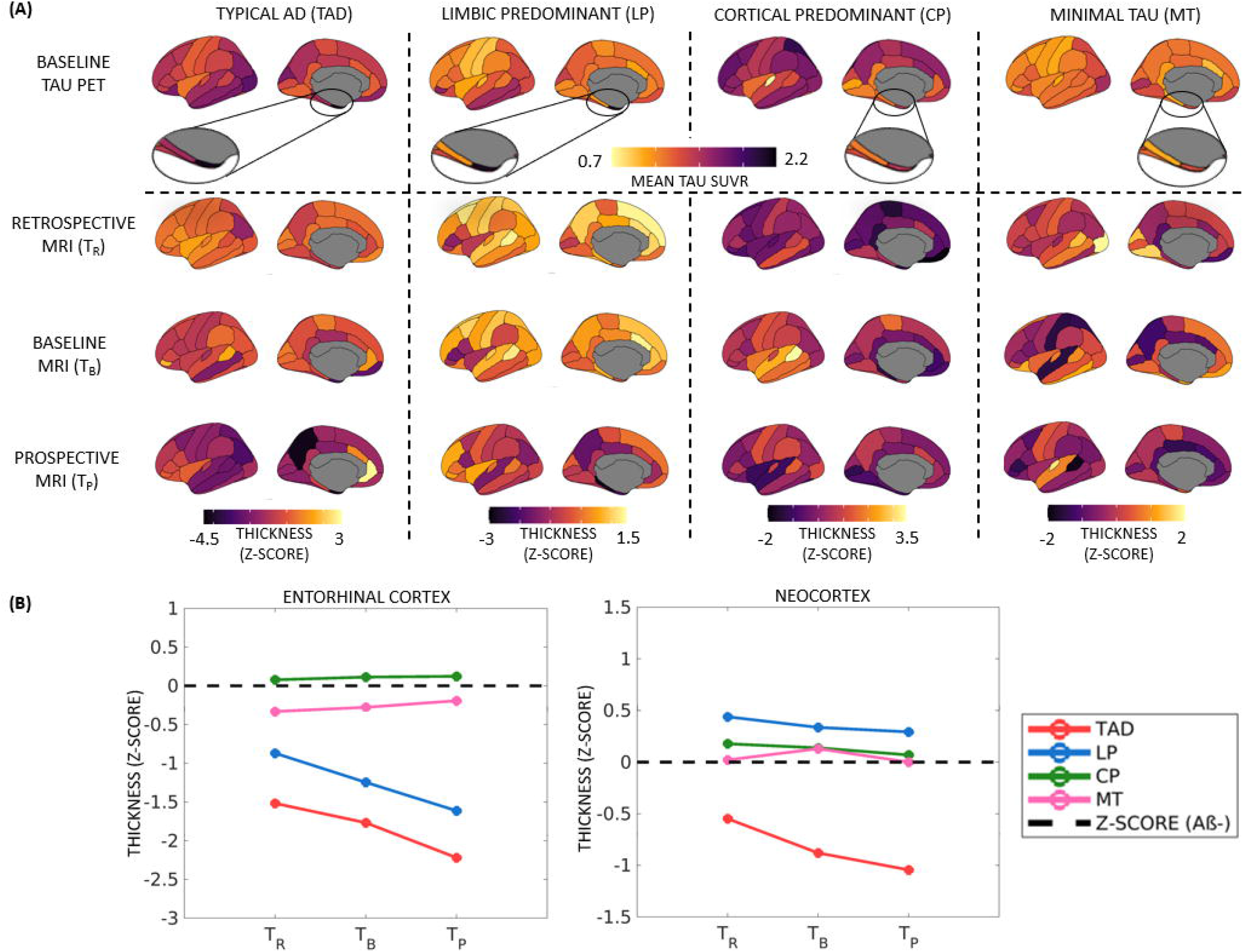
Baseline tau-PET patterns (discrete-scale) and corresponding longitudinal atrophy in the AD continuum. Note: **(A)** Top panel: Topography of baseline tau-PET SUVR in four discrete tau-PET patterns in AD continuum (N = 173). The zoomed in view shows the tau-PET SUVR in the entorhinal cortex. Darker (black) colors represent elevated tau-PET SUVR; Bottom panels: Topography of longitudinal thickness (Z-score) corresponding to each tau-PET pattern in a subcohort of AD continuum, tracked across retrospective, baseline and prospective timepoints (N = 61). Darker (black) colors represent thinner cortex. All cortical maps correspond to the left hemisphere (similar patterns were observed in the right hemisphere). **(B)** Longitudinal regional thickness (Z-score) compared across the four discrete tau-PET patterns in the entorhinal cortex and neocortex within the subcohort of AD continuum (N = 61). Z-scores were computed for each region (averaged bilaterally) relative to the Aβ-healthy group (shown in black dashed line for reference) at each timepoint. Z-scores below 0 represent regional thinning. AD = Alzheimer’s disease; T_R_ = retrospective timepoint; T_B_ = baseline timepoint; T_P_ = prospective timepoint; TAD = typical AD; LP = limbic predominant; CP = cortical predominant; MT = minimal tau; Z-SCORE (Aβ-) = reference Z-score calculated relative to the healthy reference (Aβ-) shown for reference; Aβ = amyloid-beta.

**Figure 3B** qualitatively compares longitudinal thickness *across* the tau-PET patterns in the subcohort of the AD continuum relative to the healthy (Aβ-) group. In the entorhinal cortex, all but the cortical predominant pattern showed thinning (Z-score<1) relative to the healthy group. Typical AD showed greatest thinning over time, followed by the limbic predominant pattern. Minimal tau and cortical predominant patterns showed a relatively stable entorhinal thinning over time. In the neocortex, only typical AD showed thinning/decline over time relative to the healthy (Aβ-) group.

**Table 2** presents the differential association between baseline tau-PET SUVR and longitudinal thickness change across the four tau-PET patterns. In the entorhinal cortex, typical AD pattern showed a significant association between elevated baseline tau-PET SUVR and retrospective-to-baseline thinning whereas limbic predominant pattern showed a significant association between elevated baseline tau-PET SUVR and baseline-to-prospective thinning. In the neocortex, both typical AD and cortical predominant patterns showed a significant association between elevated baseline tau-PET SUVR and baseline-to-prospective thinning.

**Table 2.** Association of baseline tau-PET and longitudinal atrophy change across tau-PET patterns (discrete scale) in the AD continuum.

|  | Entorhinal Cortex |  |  |  | Neocortex |  |  |  |
| --- | --- | --- | --- | --- | --- | --- | --- | --- |
|  | TAD | LP | CP | MT | TAD | LP | CP | MT |
| N (94) | 38 | 8 | 19 | 29 | 38 | 8 | 19 | 29 |
| Retrospective to Baseline MRI | **r=0.53, p=0.001** | r=0.42, p=0.35 | r=0.37, p=0.16 | r=-0.17, p=0.67 | r=-0.15, p=0.68 | r=-0.68, p=0.13 | r=0.51, p=0.56 | r=-0.46, p=0.11 |
| N (112) | 40 | 14 | 19 | 39 | 40 | 14 | 19 | 39 |
| Baseline to Prospective MRI | r=0.50, p=0.14 | **r=0.81, p=0.043** | r=0.37, p=0.15 | r=-0.19, p=0.44 | **r=0.59, p<0.001** | r=0.69, p=0.13 | **r=0.61, p=0.007** | r=-0.11, p=0.51 |

Significant associations are reported in **bold** in terms of r=coefficient of correlation corresponding to $p \leq 0.05$ ; All associations are adjusted for interval between timepoints. AD = Alzheimer's disease; TAD = typical AD pattern; LP = limbic predominant pattern; CP = cortical predominant pattern; MT = minimal tau pattern.

### 3.4. A/T/longitudinal-N classification in tau PET patterns

We examined the A/T/longitudinal-N biomarker scheme across the tau-PET patterns[21] (main results correspond to A assessed globally, T and longitudinal-N assessed regionally in the entorhinal cortex and neocortex; for comparison of different cutpoints, see **Supplementary Tables 5-6**). **Figure 4** and **Supplementary Table 7** show the most prevalent (observed in ≥50% of the individuals) A/T/longitudinal-N profile across tau-PET patterns: typical AD pattern was [A+ T+ N_R_+ N_B_+ N_P_+] in both the entorhinal cortex and neocortex; limbic predominant pattern was [A+ T+ N_R_+ N_B_+ N_P_+] in the entorhinal cortex but [A+ T-N_R_-N_B_+ N_P_+] in the neocortex; cortical predominant pattern was [A+ T-N_R_-N_B_-N_P_-] in the entorhinal cortex but [A+ T+ N_R_-N_B_-N_P_-] in the neocortex; minimal tau was [A+ T-N_R_-N_B_-N_P_-] in both the entorhinal cortex and neocortex.

**Figure 4.**
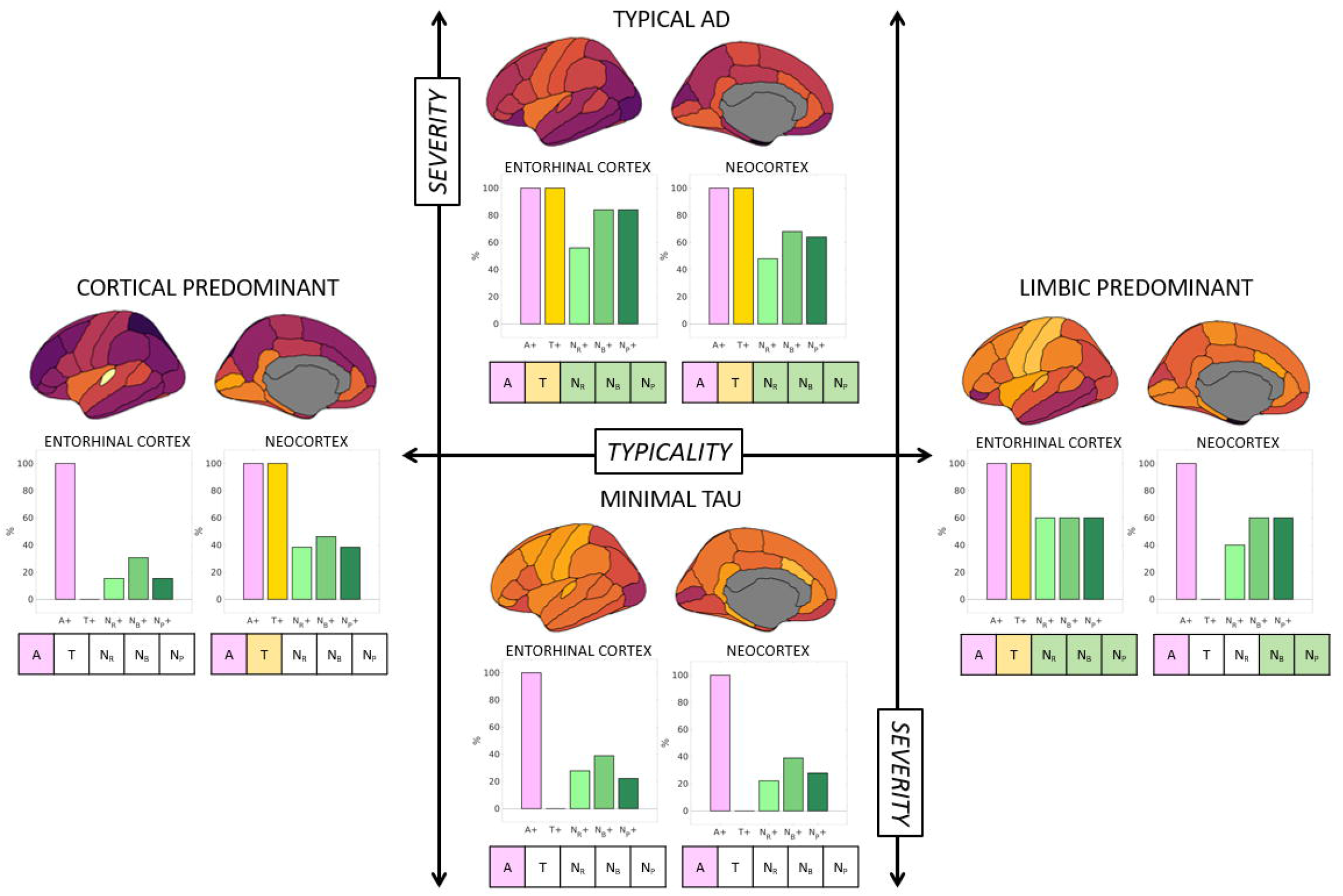
A/T/longitudinal-N classification corresponding to the tau-PET-based patterns in the AD continuum. Note: A/T/longitudinal-N biomarker profiles for the four discrete tau-PET-based patterns were mapped in the subcohort of the AD continuum (N=61). Typicality (horizontal axis) and severity (vertical axis) dimensions are superposed, as proposed in the original conceptual framework^15^. A+ was determined by global Aβ-PET SUVR. T+ and longitudinal N+ were determined in the entorhinal cortex and the neocortex, corresponding to the regions used to identify the heterogeneous tau-PET patterns^24^. For each tau-PET-based pattern, the proportion of A+, T+ and longitudinal N+ (along horizontal axis) are presented as percentages (along vertical axis) in the bar plots for the entorhinal cortex and the neocortex. Atrophy, used to represent N+, was adjusted for age at each timepoint relative to a group of healthy (Aβ-) individuals. The most prevalent A/T/longitudinal-N profile (≥50%) corresponding to a tau-PET-based pattern is shown under each bar plot with (un)colored boxes. A/T/longitudinal-N = Aβ/Tau/longitudinal-Neurodegeneration; AD = Alzheimer’s disease; N_R_ = atrophy at retrospective timepoint; N_B_ = atrophy at baseline timepoint; N_P_ = atrophy at prospective timepoint.

## 4. DISCUSSION

We investigated the association between tau-PET patterns and longitudinal neurodegeneration (atrophy) in the AD continuum. As hypothesized: (a) different tau-PET patterns revealed a differential association with longitudinal atrophy; and (b) characterizing patterns on a continuous-scale may be more sensitive than the conventional categorization of individuals into discrete patterns. Recent studies have investigated the association between tau pathology and downstream neurodegeneration in healthy, cognitively normal, prodromal AD, and AD dementia[5–8,24–26] individuals, as well as in clinical subtypes of AD[27–29]. To our knowledge, our study is the first to characterize this association relative to the biological heterogeneity[16] (tau-PET patterns) within the AD continuum.

We have, for the first time, presented a promising continuous-scale characterization of tau-PET patterns, whereby individuals were represented by typicality and severity, two continuous dimensions of biological AD subtypes proposed by the recent conceptual framework[16]. We compared it to the conventional discrete-scale characterization of heterogeneity, whereby individuals were categorized into four discrete patterns based on the contribution of the entorhinal cortex and neocortex[21]. The continuous-scale approach avoids arbitrary cutpoints, making it suitable for populations where the prevalence of different patterns are not well-known (e.g., AD continuum, beyond AD dementia) and to small cohorts. The discrete-scale approach defines patterns based on a cutpoint (e.g., Z-score>1 relative to healthy Aβ-individuals)[21,30], influencing the prevalence of the identified heterogeneous patterns. Nevertheless, both approaches share some correspondence (**Figure 1**): examining typicality, higher E:N may reflect a limbic predominant pattern while lower E:N may reflect a cortical predominant pattern; examining severity, higher global tau-PET SUVR may reflect a typical AD pattern while lower global tau-PET SUVR may reflect a minimal tau pattern. All previous subtyping methods in AD characterized heterogeneity on a discrete-scale[13–15,17], which is critical to delineate pattern-specific characteristics. However, discrete-scale characterizations often lack individual-level agreement[14]. A continuous-scale characterization of heterogeneity may be more sensitive, free from the assumption of pre-defined prevalence in a population. Hence, we encourage future studies to explore and validate new operationalizations of typicality and severity representing disease heterogeneity.

The four tau-PET patterns captured by the continuous- and discrete-scales in the AD continuum in our study are reminiscent of the biological tau-PET AD subtypes[13,14,17,31]. Heterogeneous tau-PET topographies which may represent current *patterns* in our cohort at early disease stage, may be more appropriately described as *subtypes* in AD dementia. Our study confirms the findings from the recent report identifying four discrete trajectories in tau-PET within AD continuum[15]. The novelty of our findings lies in the associations between baseline tau-PET and longitudinal atrophy across heterogeneity and the notion of heterogeneity being a continuous phenomenon.

The prevalence of the identified tau-PET patterns differed slightly from previous reports: on the continuous-scale (**Figure 1A-B**), a large proportion of the individuals exhibited intermediate values of typicality and low values of severity (lower variance in prodromal AD and cognitively normal may suggest lesser heterogeneity); on the discrete-scale (**Figure 3A**), minimal tau was the most prevalent pattern (37%). These findings are likely owing to the large proportion of individuals at early disease stages (cognitively normal and prodromal AD), who may have not accumulated considerable amount of tau pathology, typical to AD. Additionally, a current tau-PET pattern at early disease stages may likely evolve into a different pattern later. This may explain why the demographic/clinical profiles of our tau-PET patterns (**Supplementary Table 2**) do not entirely conform with the expected profiles previously reported in AD[16]. Similar results have been found when characterizing heterogeneity in tau-PET in the AD continuum[15], atrophy in prodromal AD[32] and glucose-hypometabolism in prodromal AD[33]. Altogether, tau-PET patterns at preclinical and prodromal stages of AD may be similar to, albeit less pronounced than, heterogeneity in AD dementia.

Our main finding was that tau-PET patterns showed differential association with longitudinal atrophy. On the continuous-scale (**Supplementary Table 3**), typicality was associated with retrospective-to-baseline atrophy whereas severity was additionally associated with baseline-to-prospective atrophy. This may suggest that typicality-related atrophy changes may precede severity-related atrophy changes. Significant associations were stronger for the entorhinal cortex than the neocortex. Additionally, retrospective-to-baseline atrophy was significant relative to typicality and severity (**Figure 2**). Altogether, these highlight that the highest extreme of typicality (limbic predominant pattern) may reflect greater and earlier vulnerability of the entorhinal cortex than neocortex at the early stages of AD[34], while the lowest extreme of typicality (cortical predominant pattern) may reflect a low selective vulnerability or increased resilience of the entorhinal cortex within a subgroup of the AD continuum. Severity may better capture atrophy-based neurodegeneration, not limited to AD pathology alone[1]. Lack of significant prospective atrophy changes (**Figure 2**) could potentially be attributed to a shorter baseline-to-prospective interval (1.3±0.4years) than the retrospective-to-baseline interval (2.2±1years), not allowing us to observe potential significant atrophy changes prospectively.

On the discrete-scale (**Table 2**), baseline tau-PET patterns were associated with longitudinal atrophy change for typical AD and limbic predominant patterns in the entorhinal cortex but for typical AD and cortical predominant patterns in the neocortex. This result highlights a region-specific differential association between tau-PET patterns and atrophy. Typical AD and limbic predominant patterns showed topographical correspondence between tau-PET and (prospective) MRI while cortical predominant and minimal tau did not (**Figure 3**). The two latter patterns showed marked atrophy in brain regions non-specific to the tau-PET patterns (entorhinal atrophy in cortical predominant; cortical atrophy in minimal tau), indicating that atrophy may not always regionally follow the different tau-PET patterns. Conversely, topographical correspondence has been reported between tau-PET and MRI in atrophy-based AD subtypes[35]. Together, these findings may imply that heterogeneity of a downstream event (atrophy) may be reflected in an upstream event (tau pathology) but not vice-versa. Moreover, the downstream contributions of other neuropathologies towards atrophy need to be considered in the future as biomarkers for those pathologies become available.

Across the continuous- and discrete-scale characterizations of tau-PET patterns, baseline-to-prospective atrophy associated with baseline tau pathology supports the hypothesis of tau as a possible driver of atrophy[4,6,24,36], observed across some but not necessarily all tau-PET patterns. However, we also found retrospective-to-baseline atrophy associated with baseline tau pathology in some but not all tau-PET patterns, suggesting possible non-linear atrophy trajectories across tau-PET patterns.

Furthermore, we demonstrated differential profiles of the A/T/longitudinal-N biomarker scheme across tau-PET patterns (**Figure 4**). While the limbic predominant pattern demonstrated T+ in the entorhinal cortex and T- in the neocortex, the cortical predominant pattern demonstrated the opposite profile. This contrast may suggest a non-uniform sequence of tau accumulation across the tau-PET patterns. This aligns with the proposed hypothesis of alternative possible pathways for initiation/spread of tau pathology in the cortical-predominant pattern [37]. All patterns showed some longitudinal neurodegeneration (adjusted for age), but only typical AD and limbic predominant patterns showed ≥50% prevalence of longitudinal N+. Combined with reports suggesting a preferential association of atrophy to tau pathology over Aβ[4,38], this result may imply that atrophy may not entirely be tau-related and could be partly tau-independent, extending beyond the effect of normal aging. One caveat, however, is that the prevalence of A/T/longitudinal-N profiles varied widely depending on the cutpoint used (**Supplementary Tables 5-7**), an issue that is known in the field[1] which should be taken into account in future studies.

Our study has some limitations. Although the overall goal of our study was to understand the heterogeneity in tau-PET patterns across the AD continuum, cognitively normal individuals (Aβ+) were overrepresented. This dominance of the early stages of AD likely translated to the relatively less pronounced tau-PET patterns. Moreover, the ability of [^18^F]AV-1451 tracer in detecting tau pathology may be limited at these early disease stages[39]. Hippocampus, a key region in most neuropathological and MRI studies investigating heterogeneity in AD[11,12,21,40– 42], was not evaluated as its signal is confounded by off-target binding in tau-PET[19,20]. However, we have previously shown that tau-PET-based heterogeneous patterns based on the entorhinal cortex are similar to those based on hippocampus[14]. Although we tracked the longitudinal atrophy changes relative to baseline tau pathology, we could not assess longitudinal tau-PET changes. Finally, considering the strict inclusion criteria in ADNI, the generalizability of our findings in a heterogenous/clinical setting remains to be validated.

In conclusion, we demonstrated that the associations are not the same between different tau-PET patterns and longitudinal atrophy in the AD continuum. Secondarily, we posit treating heterogeneity in the AD continuum as a continuous phenomenon over the conventional discrete categorization. Together, our findings can have practical implications towards the design of clinical trials, development of targeted therapeutics and ultimately, realization of precision medicine.

## Data Availability

All data used in this work is publicly available for registered users: http://adni.loni.usc.edu/

## ACKNOWLEDGEMENTS

This study was funded by the Swedish Foundation for Strategic Research (SSF); the Strategic Research Programme in Neuroscience at Karolinska Institutet (StratNeuro); the Swedish Research Council (VR); the regional agreement on medical training and clinical research (ALF) between Stockholm County Council and Karolinska Institutet; Center for Innovative Medicine (CIMED); the Swedish Alzheimer Foundation; the Swedish Brain Foundation; the Åke Wiberg Foundation; Demensfonden; Stiftelsen Olle Engkvist Byggmästare; Birgitta och Sten Westerberg; Foundation for Geriatric Diseases at Karolinska Institutet; Loo och Hans Ostermans stiftelse för medicinsk forskning; Stiftelsen För Gamla Tjänarinnor; Gun & Bertil Stohnes Stiftelse. The funding sources did not have any involvement in the study design; collection, analysis, and interpretation of data; writing of the report; and the decision to submit the article for publication.

Data collection and sharing for this study was funded by the Alzheimer’s Disease Neuroimaging Initiative (ADNI) (National Institutes of Health Grant U01 AG024904) and DOD ADNI (Department of Defense award number W81XWH-12-2-0012). ADNI is funded by the National Institute on Aging, the National Institute of Biomedical Imaging and Bioengineering, and through generous contributions from the following: Alzheimer’s Association; Alzheimer’s Drug Discovery Foundation; BioClinica, Inc.; Biogen Idec Inc.; Bristol-Myers Squibb Company; Eisai Inc.; Elan Pharmaceuticals, Inc.; Eli Lilly and Company; F. Hoffmann-La Roche Ltd and its affiliated company Genentech, Inc.; GE Healthcare; Innogenetics, N.V.; IXICO Ltd.; Janssen Alzheimer Immunotherapy Research & Development, LLC.; Johnson & Johnson Pharmaceutical Research & Development LLC.; Medpace, Inc.; Merck & Co., Inc.; Meso Scale Diagnostics, LLC.; NeuroRx Research; Novartis Pharmaceuticals Corporation; Pfizer Inc.; Piramal Imaging; Servier; Synarc Inc.; and Takeda Pharmaceutical Company. The Canadian Institutes of Health Research is providing funds to support ADNI clinical sites in Canada. Private sector contributions are facilitated by the Foundation for the National Institutes of Health (www.fnih.org). The grantee organization is the Northern California Institute for Research and Education, and the study is coordinated by the Alzheimer’s Disease Cooperative Study at the University of California, San Diego. ADNI data are disseminated by the Laboratory for Neuro Imaging at the University of California, Los Angeles.

Data used in this study were obtained from the Alzheimer’s Disease Neuroimaging Initiative (ADNI) database (adni.loni.ucla.edu). As such, the investigators within the ADNI contributed to the design and implementation of ADNI and/or provided data but did not participate in the analysis or writing of this report. A complete listing of ADNI investigators can be found at: http://adni.loni.ucla.edu/wp-content/uploads/how_to_apply/ADNI_Acknowledgement_List.pdf.

## Notes

### Competing Interest Statement

The authors have declared no competing interest.

### Author Declarations

Data used in this work comes from publicly available dataset, the ADNI. All protocols for the ADNI were approved by the local institutional review boards of the participating sites.

