## Supplementary Document for "Associations between different tau-PET patterns and longitudinal atrophy in the Alzheimer’s disease continuum"

**Supplementary Material**

### 1. Participants

The selected cohort (N=366) comprised individuals on the AD continuum who were Aβ+, reflecting presence of AD pathology (N=173) and cognitively normal healthy individuals who were Aβ-, reflecting absence of preclinical AD pathology (N=193). The detailed inclusion and exclusion criteria for the ADNI can be found at http://adni.loni.usc.edu/methods/. The interval between tau-PET and MRI at baseline was about 90 days. Aβ status was determined through amyloid PET (florbetapir SUVR cut-off = 1.11 [1]; or florbetaben SUVR cut-off = 1.08 from http://adni.loni.usc.edu/).

### 2. Neuroimaging processing

*MRI processing*

The MRI were processed through TheHiveDB system [2] using FreeSurfer 6.0.0 (http://freesurfer.net/). Data were first preprocessed through the cross-sectional FreeSurfer stream. Resulting segmentations were visually screened for quality control. Screened scans were included for further preprocessing through the longitudinal FreeSurfer stream [3]. Automatic region of interest parcellation yielded thickness in 68 cortical areas [4], serving as a measure of brain atrophy.

*Tau-PET processing*

Tau-PET scans were processed using the PetSurfer Toolbox [5] within FreeSurfer 6.0.0. AV-1451 images were co-registered onto the cross-sectionally processed MRI and visually assessed for alignment. Estimated regions of interest were consistent with those used for MRI [4]. We chose to perform and report only partial volume corrected (PVC) values using the symmetric geometric matrix method [6] based on a previous subtyping study which demonstrated reasonable agreement between PVC and non-PVC data in this cohort [7]. Regional AV-1451 signal was quantified in terms of the standardized uptake value ratio (SUVR), computed with the cerebellum grey matter as the reference.

**3. Subtyping algorithm to characterize heterogeneity in tau-PET within the AD continuum on a discrete scale** [8]

*Measure:* cortical tau-PET SUVR

*Regions:* medial temporal lobe represented by entorhinal cortex; neocortex represented by middle frontal, superior temporal, inferior parietal regions; each region averaged across both cerebral hemispheres

*Method:*

(i) Z-score normalization: All regional tau-PET SUVR values were converted into Z-scores by comparing against the healthy (Aβ-) reference group.

(ii) Abnormality Identification: Z-scores of tau-PET SUVR in the entorhinal cortex ($Z_E$), and neocortex (average of frontal, temporal, parietal regions, $Z_N$) of individuals were calculated based on the normative data. A cutpoint of Z-score>1.0 determined the presence of abnormal tau pathology in each ROI.

(iii) Classification: Four discrete patterns were identified. If $Z_E>1$ and $Z_N>1$, then individuals were classified as the *typical AD* pattern. If $Z_E>1$ and $Z_N\leq1$, then individuals were classified as the *limbic predominant* pattern. If $Z_E\leq1$ and $Z_N>1$, then individuals were classified as *cortical predominant* pattern. If $Z_E\leq1$ and $Z_N\leq1$, then individuals were classified as the *minimal tau* pattern.

**4. Association between baseline tau-PET and longitudinal atrophy change for tau-PET heterogeneity (continuous scale) in the AD continuum**

*Linear regression model*

Independent variable was baseline regional tau-PET SUVR. Dependent variable was regional thickness change between two timepoints. The interval between the two timepoints was used as a covariate. Regional thickness change between two timepoints was evaluated as follows:

$$thickness\ change\ = \left(\frac{thickness\ at\ timepoint\ 1\ -\ thickness\ at\ timepoint\ 2}{thickness\ at\ timepoint\ 1}\right)$$

*Linear mixed effects model*

We used individual-specific intercepts and the fixed effects included time (centered at baseline, $T_B$), age, baseline regional tau-PET SUVR, typicality, severity, interaction of each of baseline regional tau-PET SUVR, typicality, severity with time. Dependent variable was regional thickness.

**5. Tau positivity in tau-PET patterns in the AD continuum**

(i) *Byun's cutpoint:* tau positivity was defined as the Z-score of tau-PET SUVR > 1 relative to the healthy (Aβ-) reference group, i.e., $Z_E>1$ for the entorhinal cortex and $Z_N>1$ for the neocortex (middle frontal, superior temporal, inferior parietal) based on a previous subtyping study[8].

(iii) *Braak staging-based cutpoint:* tau positivity was defined for the entorhinal (Braak stage I) as PVC tau SUVR > 1.129 (the hippocampus corresponding to Braak stage II was excluded due to potential off-target binding in the region) and for cortical brain areas including frontal cortex, parietal cortex, occipital cortex, transverse, superior temporal cortex, precuneus, banks of superior temporal sulcus, precentral gyrus, postcentral gyrus, paracentral gyrus, cuneus, pericalcarine (Braak stages V-VI) as PVC tau SUVR > 1.873 based on a previous tau-PET study[9].

Consistent with our regions of interest (entorhinal cortex and neocortex), our main findings correspond to the results from Byun's cutpoints above.

**6. Neurodegeneration positivity in tau-PET patterns in the AD continuum**

(i) *Byun's cutpoint:* neurodegeneration positivity was defined as the Z-score of MRI-based thickness < 1 relative to the healthy (Aβ-) reference group, i.e., $Z_E<1$ for the entorhinal cortex and $Z_N<1$ for the neocortex (middle frontal, superior temporal, inferior parietal) based on a previous subtyping study[8].

(ii) *Meta ROI-based cutpoint:* neurodegeneration positivity was defined in a meta ROI in the medial temporal lobe (entorhinal, inferior temporal, middle temporal, fusiform) as thickness < 2.67 mm based on a previous study establishing imaging-based cutpoints in AD[10]. Due to unavailability of a specific threshold for the cortex, the same threshold of 2.67 mm was applied to the neocortex (middle frontal, superior temporal, inferior parietal).

Consistent with our regions of interest (entorhinal cortex and neocortex), our main findings correspond to the results from Byun's cutpoints above.

**Table 1.** Characteristics of the study cohort

| Diagnosis (baseline) | Healthy (Aβ-) | Cognitively Normal (Aβ +) | Prodromal AD (Aβ +) | AD Dementia (Aβ +) |
|---|---|---|---|---|
| **N (baseline tau-PET, MRI)** | 193 | 98 | 50 | 25 |
| **N (retrospective MRI)** | 73 | 46 | 31 | 17 |
| **N (prospective MRI)** | 66 | 57 | 42 | 13 |
| **Age at baseline (years)** | 71.9 ± 6.4 [56 95] | 75.5 ± 7.1 [62 92][a] | 75.3 ± 7.7 [59 92][a] | 78.2 ± 8.2 [56 91][a] |
| **Sex (% Female)** | 59.6 | 56.1 | 52 | 48 |
| **Education (years)** | 17 ± 2.3 [11 20][b] | 16.7 ± 2.3 [12 20][b] | 15.6 ± 2.6 [12 20] | 15.9 ± 2.6 [12 20] |
| ***APOE* ε4 carriers (%)[e]** | 23.6 | 56.7[a] | 62[a] | 56[a] |
| **MMSE at baseline[f]** | 29.3 ± 1.0 [23 30][c] | 28.8 ± 1.5 [22 30][c] | 27.6 ± 2.3 [19 30][d] | 22 ± 4.2 [9 30][d] |

Data are reported as mean ± standard deviation [minimum, maximum]; Hypothesis testing was performed using the Kruskal–Wallis test for the continuous variables and Fisher exact test for the nominal variables. [a] significantly different from Healthy (Aβ-); [b] significantly different from Prodromal AD; [c] significantly different from Prodromal AD and AD Dementia; [d] significantly different from all other groups; [e] Missing values = 3; [f] Missing values = 2; Aβ = β-amyloid; AD = Alzheimer's disease; *APOE* = apolipoprotein; MMSE = mini mental state examination.

**Table 2.** Characteristics of the tau-PET patterns (discrete scale) in the AD continuum

| AD continuum at baseline (N=173) | | | | |
|---|---|---|---|---|
| | **TAD (N=57)** | **LP (N=21)** | **CP (N=31)** | **MT (N=64)** |
| **Age at baseline (years)** | 76.3 ± 7.3 [62, 90] | 76.3 ± 6.4 [65, 90] | 76.6 ± 7.8 [56, 91] | 74.9 ± 7.8 [59, 92] |
| **Sex (% Female)** | 64.9 | 42.9 | 54.8 | 46.9 |
| **Education (years)** | 15.9 ± 2.3 [12, 20][a] | 15.6 ± 2.6 [12, 20][a] | 16.1 ± 2.4 [12, 20] | 17 ± 2.4 [12, 20] |
| ***APOE* ε4 carriers (%)** | 66.7 | 57.1 | 58.1 | 50.8 |
| **MMSE** | 26.1 ± 4.3 [9, 30][a] | 26.8 ± 3.1 [20, 30][a, b] | 28.2 ± 2.1 [21, 30] | 28.6 ± 1.9 [17, 30] |
| **ADNI-MEM** | 0.10 ± 0.96 [-2.38, 2.01][a, b] | 0.19 ± 0.62 [-1.19, 1.21][a] | 0.55 ± 0.66 [-0.84, 1.69][a] | 0.93 ± 0.56 [-0.40, 2.10] |
| **ADNI-EF** | 0.04 ± 1.12 [-2.43, 2.99][a, c] | 0.62 ± 1.01 [-1.40, 2.00] | 0.43 ± 0.88 [-2.78, 2.23][a] | 0.94 ± 0.87 [-2.31, 2.72] |

Data are reported as mean ± standard deviation [minimum, maximum]; Hypothesis testing was performed using the Kruskal–Wallis test for the continuous variables and Fisher exact test for the nominal variables. [a] significantly different from minimal tau pattern; [b] significantly different from cortical predominant pattern; [c] significantly different from limbic predominant pattern; AD=Alzheimer's disease; TAD=typical AD pattern; LP=limbic predominant pattern; CP=cortical predominant pattern; MT=minimal tau pattern; *APOE*=apolipoprotein; MMSE=mini mental state examination; ADNI-MEM=composite cognitive scores for memory; ADNI-EF= composite cognitive scores for executive function.

**Table 3.** Association of baseline tau-PET and change in atrophy for tau-PET heterogeneity (continuous scale) in the AD continuum

| | | Baseline Typicality (tau-PET) vs Change in Thickness (MRI) | | Baseline Severity (tau-PET) vs Change in Thickness (MRI) | |
|---|---|---|---|---|---|
| **Time** | **Group** | **Entorhinal Cortex** | **Neocortex** | **Entorhinal Cortex** | **Neocortex** |
| Retrospective to Baseline MRI | Aβ+ (N = 94) | **r=0.43, *p*<0.001** | **r=0.23, *p*=0.02** | **r=0.46, *p*<0.001** | r=0.15, *p*=0.2 |
| | Aβ- (N = 73) | r=0.10, *p*=0.73 | r=-0.15, *p*=0.29 | r=0.21, *p*=0.14 | r=0.13, *p*=0.61 |
| Baseline to Prospective MRI | Aβ+ (N = 112) | r=0.18, *p*=0.10 | r=-0.20, *p*=0.05 | **r=0.39, *p*<0.001** | **r=0.35, *p*<0.001** |
| | Aβ- (N = 66) | r=0.02, *p*=0.91 | r=-0.03, *p*=0.82 | r=0.21, *p*=0.98 | r=-0.21, *p*=0.90 |

Significant associations in terms of r=coefficient of correlation and corresponding to *p*≤0.05 are reported in **bold**. Change in thickness from timepoint 1 to timepoint 2 $= \left(\frac{thickness\ at\ timepoint\ 1 - thickness\ at\ timepoint\ 2}{thickness\ at\ timepoint\ 1}\right)$; All associations are adjusted for interval between timepoints; Aβ = β-amyloid.

**Table 4.** Distribution of the clinical groups across the tau-PET patterns (discrete-scale) in the AD continuum

| **AD continuum including all possible longitudinal MRI (baseline N = 173)** | | | | |
|---|---|---|---|---|
| **Clinical Group (Aβ+)** | **TAD (N = 57)** | **LP (N = 21)** | **CP (N = 31)** | **MT (N = 64)** |
| Cognitively Normal (%) | 30 | 52 | 61 | 80 |
| Prodromal AD (%) | 42 | 24 | 32 | 17 |
| AD Dementia (%) | 28 | 24 | 6 | 3 |
| **Subcohort of AD continuum including longitudinal MRI at all three timepoints (N = 61)** | | | | |
| **Clinical Group (Aβ+)** | **TAD (N = 25)** | **LP (N = 5)** | **CP (N = 13)** | **MT (N = 18)** |
| Cognitively Normal (%) | 32 | 40 | 46 | 72 |
| Prodromal AD (%) | 48 | 20 | 54 | 28 |
| AD Dementia (%) | 20 | 40 | 0 | 0 |

Percentages are rounded to nearest integer. AD=Alzheimer's disease; Aβ=amyloid-beta; TAD=typical AD pattern; LP=limbic predominant pattern; CP=cortical predominant pattern; MT=minimal tau pattern.

**Table 5.** Comparison of cutpoints to determine baseline tau positivity in tau-PET patterns in the AD continuum

| AD continuum (Aβ+) at baseline (N=173) | | | | | |
|---|---|---|---|---|---|
| **Region** | **Method** | **TAD (N=57)** | **LP (N=21)** | **CP (N=31)** | **MT (N=64)** |
| **T+ % MTL** | Byun | 100 | 100 | 0 | 0 |
| | Braak | 100 | 100 | 100 | 93.7 |
| **T+ % Cortex** | Byun | 100 | 0 | 100 | 0 |
| | Braak | 85.9 | 14.3 | 83.9 | 3.12 |

T+=tau positivity; AD=Alzheimer's disease; Aβ=amyloid-beta; TAD=typical AD pattern; LP=limbic predominant pattern; CP=cortical predominant pattern; MT=minimal tau pattern; MTL=representation of the medial temporal lobe.

**Table 6.** Comparison of cutpoints to determine longitudinal neurodegeneration positivity in tau-PET patterns in the AD continuum

| **Subcohort of AD continuum (Aβ+) with longitudinal MRI at all three timepoints (N=61)** | | | | | | | | | | | | | |
|---|---|---|---|---|---|---|---|---|---|---|---|---|---|
| | | **TAD (N=38)** | | | **LP (N=8)** | | | **CP (N=19)** | | | **MT (N=29)** | | |
| **Region** | **Method** | $T_R$ | $T_B$ | $T_P$ | $T_R$ | $T_B$ | $T_P$ | $T_R$ | $T_B$ | $T_P$ | $T_R$ | $T_B$ | $T_P$ |
| **N+ % MTL** | Byun | 56 | 84 | 84 | 60 | 60 | 60 | 15.4 | 30.8 | 15.4 | 27.8 | 38.9 | 22.2 |
| | Meta ROI | 40 | 68 | 68 | 20 | 60 | 60 | 15.4 | 30.8 | 23.1 | 11.1 | 16.7 | 11.1 |
| **N+ % Cortex** | Byun | 48 | 68 | 64 | 40 | 60 | 60 | 38.5 | 46.1 | 38.5 | 22.2 | 38.9 | 27.8 |
| | Meta ROI | 80 | 96 | 96 | 100 | 100 | 100 | 76.9 | 84.6 | 84.6 | 77.8 | 88.9 | 88.9 |

N+=neurodegeneration positivity; N+ values are adjusted for age at each timepoint; AD=Alzheimer's disease; Aβ=amyloid-beta; TAD=typical AD pattern; LP=limbic predominant pattern; CP=cortical predominant pattern; MT=minimal tau pattern; MTL=representation of the medial temporal lobe; $T_R$=retrospective timepoint; $T_B$=baseline timepoint; $T_P$=prospective timepoint.

**Table 7.** A/T/longitudinal-N biomarker scheme corresponding to the tau-PET patterns in the AD continuum

| **AD continuum (100% A+) at baseline (N=173)** | | | | | |
|---|---|---|---|---|---|
| | | **TAD (N=57)** | **LP (N=21)** | **CP (N=31)** | **MT (N=64)** |
| **Entorhinal Cortex** | **T+ %** | 100 | 100 | 0 | 0 |
| | **N+ %** | 63.2 | 52.4 | 29.0 | 28.1 |
| **Neocortex** | **T+ %** | 100 | 0 | 100 | 0 |
| | **N+ %** | 50.9 | 42.9 | 41.9 | 26.6 |
| **Subcohort of AD continuum (100% A+) with longitudinal MRI at all three timepoints (N=61)** | | | | | |
| | | **TAD (N=38)** | **LP (N=8)** | **CP (N=19)** | **MT (N=29)** |
| **N+ % Entorhinal Cortex** | $T_R$ | 56 | 60 | 15.4 | 27.8 |
| | $T_B$ | 84 | 60 | 30.8 | 38.9 |
| | $T_P$ | 84 | 60 | 15.4 | 22.2 |
| **N+ % Neocortex** | $T_R$ | 48 | 40 | 38.5 | 22.2 |
| | $T_B$ | 68 | 60 | 46.1 | 38.9 |
| | $T_P$ | 64 | 60 | 38.5 | 27.8 |

A+=Aβ positivity with global PET Aβ SUVR; T+=tau positivity with Byun's cutpoint; N+=neurodegeneration positivity with Byun's cutpoint; N+ values are adjusted for age at each timepoint; AD=Alzheimer's disease; TAD=typical AD pattern; LP=limbic predominant pattern; CP=cortical predominant pattern; MT=minimal tau pattern.
